# Algorithmic discovery of dynamic models from infectious disease data

**DOI:** 10.1101/19012724

**Authors:** Jonathan Horrocks, Chris T. Bauch

## Abstract

Theoretical models are typically developed through a deductive process where a researcher formulates a system of dynamic equations from hypothesized mechanisms. Recent advances in algorithmic methods can discover dynamic models inductively– directly from data. Most previous research has tested these methods by rediscovering models from synthetic data generated by the already known model. Here we apply Sparse Identification of Nonlinear Dynamics (SINDy) to discover mechanistic equations for disease dynamics from case notification data for measles, chickenpox, and rubella. The discovered models provide a good qualitative fit to the observed dynamics for all three diseases, However, the SINDy chickenpox model appears to overfit the empirical data, and recovering qualitatively correct rubella dynamics requires using power spectral density in the goodness-of-fit criterion. When SINDy uses a library of second-order functions, the discovered models tend to include mass action incidence and a seasonally varying transmission rate–a common feature of existing epidemiological models for childhood infectious diseases. We also find that the SINDy measles model is capable of out-of-sample prediction of a dynamical regime shift in measles case notification data. These results demonstrate the potential for algorithmic model discovery to enrich scientific understanding by providing a complementary approach to developing theoretical models.

## Introduction

Dynamic models that capture the governing mechanisms of systems lie at the heart of many scientific theories of natural systems, ranging from planetary motion to epidemiology^1^. Since Isaac Newton’s *Principia mathematica*, research have been devoted to creating models that accurately describe and predict the behaviour of these systems^2^. These models are typically arrived at through a deductive process by hypothesizing mechanisms, formulating dynamic mathematical models that represent those mechanisms, and testing them against data. This tried-and-true approach remains essentially unchanged today.

In the late twentieth century, methods of reconstructing phase spaces or differential equation models from time series data were proposed^3,4^. More recently, advances in algorithmic sophistication, computational power, and increased data availability have renewed the development of ‘model discovery’ methods for dynamical system that determine a system of governing dynamical equations from a given dataset^5^. A seminal paper in model discovery uses symbolic regression to recover nonlinear differential equations^6^. This approach automated the process of finding the symbolic structure of the dynamical system governing a natural process. Being able to model a system symbolically rather than numerically is crucial due to the explanatory value of a model built with elementary functions, since in principle it allows prediction under a broad range of possible conditions and not just a replication of the given dataset. In other words, the technique is intended to automatically uncover the nonlinearities that govern system dynamics.

However, early attempts at dynamical systems model discovery were subject to overfitting, as well as being computationally expensive and lacking the ability to scale well to systems with higher dimensionality. Deriving dynamic models from data faces several challenges stemming from large dimensionality. The simplest method to obtain a model that explains the data well minimizes the residual squared error between the predicted response and the data (OLS). This tends to create very complicated models with high descriptive value. However, the models tend to be overfitted to any noise present in the data, compromising their predictive ability^7^. Highly complicated models also exceed human analytic ability, thus detracting from the interpretability of the model.

Most alternative methods to OLS either use subset selection, which attempts to identify some subset of the predictors that adequately describes the system while disregarding the rest^8,9^, or shrinkage (regularization), which fits the model using all of the available predictors but forces the coefficients of select predictors towards zero, thereby performing a kind of variable selection. For instance, SINDy (Sparse Identification of Nonlinear Dynamics) is a recent breakthrough that automates model discovery through sparsity-promoting regression techniques^10–18,32^. SINDy begins with a large library of nonlinear terms. The algorithm fits the model to the data with the current library, removes any nonlinear terms from the library that have small fitted coefficients, and repeats the process. This progressively shrinks the size of the library until a relatively small system of differential or difference equations with good explanatory power for the given dataset is obtained. This inductive approach contrasts with the deductive approach of first formulating a model, inferring its parameters from data, and then testing its predictive power^19^.

Epidemiological systems have long been studied with dynamic models, on account both of their public health relevance and the complex patterns exhibited by epidemics (Figure 1)^20–27^. Contemporary epidemic modelling approaches can be traced to the work of Kermack and McKendrick in 1927^28^. Their compartmental model partitions a population into a series of mutually exclusive compartments–such as susceptible, infected or recovered–and models how individuals move between the compartments^22,29,30^. Nonlinearity typically arises from infection transmission. For instance, the commonly used massaction mixing principle posits that the number of new infections is the product of the number of susceptible and infectious individuals^22,29,30^. This principle was originally used to describe the rate of a well-mixed chemical reaction by relating it to the concentration of reactants^31^, hence compartmental models conceptualize new infections as resulting from random mixing between susceptible and infectious individuals. These models have been expanded in many ways to account for observed epidemic patterns. For instance, seasonal variation in the transmission rate can give rise to complex dynamics such as bifurcations, oscillations and deterministic chaos^20–27^. The spatial and temporal dynamics of many childhood infectious disease such as measles, pertussis, rubella and chickenpox are well-described by these dynamic models.

**Figure 1.**
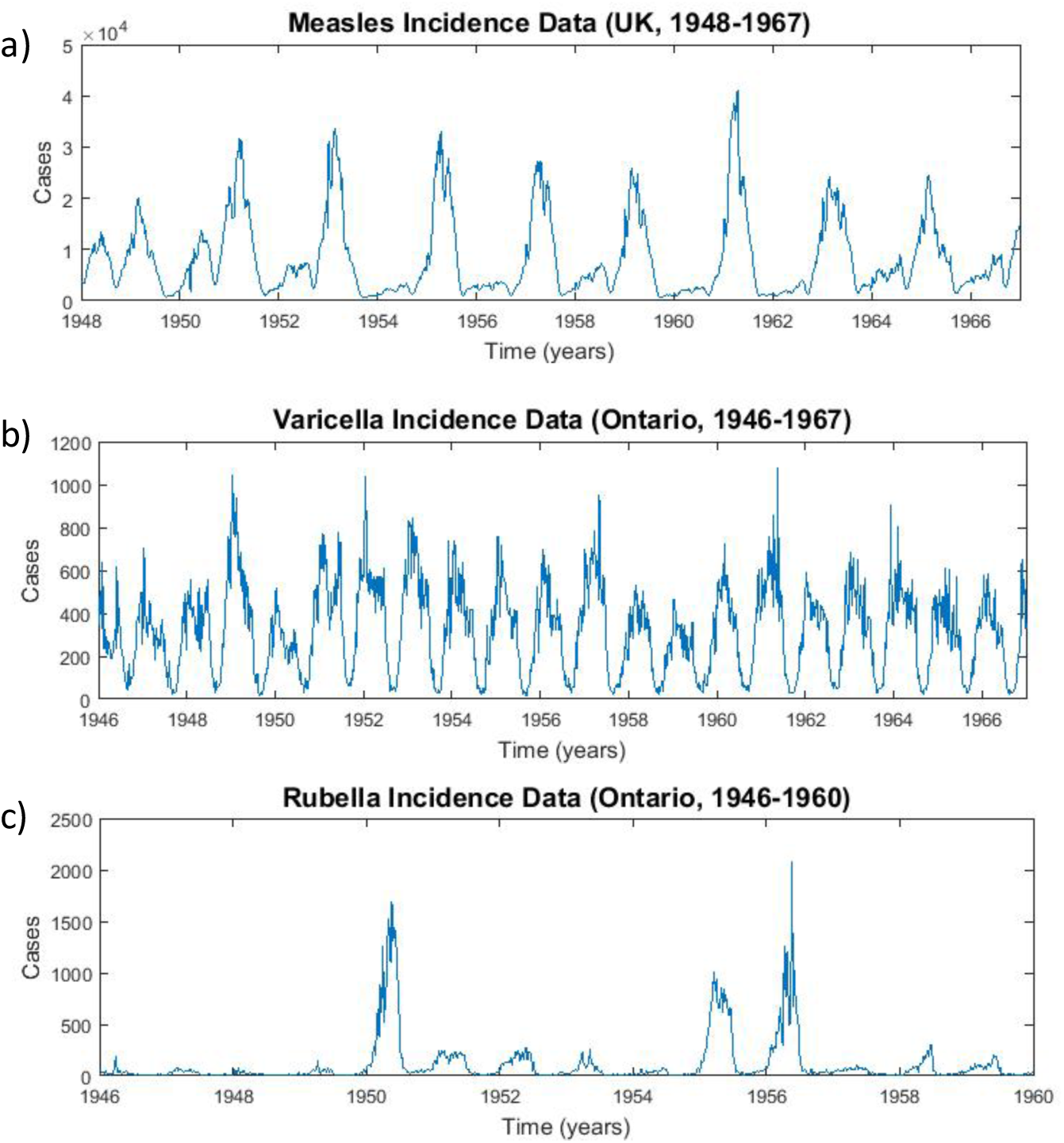
Case notifications (number of cases reported each week) for (a) measles in the United Kingdom, (b) chickenpox in Ontario and (c) rubella in Ontario. Data are from the International Infectious Disease Data Archive (http://iidda.mcmaster.ca/).

Epidemic models are one of several model systems that have been used to validate SINDy through an approach known as model rediscovery^11,32^. In this approach, synthetic data are generated by adding noise to the output from a pre-specified model, and the algorithm is applied to the synthetic data to study the conditions under which it can discover the original model^10^. Through this process, SINDy has shown it can not only successfully rediscover the original epidemic models, but it many cases it can also generate hierarchies of models of varying complexity, including new models that can fit the synthetic data but have different model equations from the original model^11,32^. SINDy has also been used for discovery of new models from empirical data in experimental mechanics and optics^33,34^ as well as to discovery of new models and model reductions from synthetic fluid mechanics data^35,36^. Thus far, however, SINDy has not yet been tested in model discovery from empirical data on infectious disease dynamics. This represents a very different challenge for model discovery algorithms: the complicated and noisy nature of real-world epidemiological data represents a challenge for generating sparse, low-dimensional models that can provide mechanistic insights or predict real-world dynamics.

Here, using minimal prior epidemiological knowledge, we apply SINDy to time series data from measles, rubella and chickenpox to discover dynamic models that govern their epidemic patterns, and we compare the resulting models to a standard compartmental model for these infections. We choose epidemiological systems because complex biological systems present a nontrivial challenge for SINDy, and these three infectious diseases display a diverse range of nontrivial dynamical behaviour. At the same time, a significant amount of research in compartmental epidemic models suggests that we have a good idea of what kind of discovered models would satisfy the criteria of interpretability and predictive power we expect from a dynamic model. This exercise allows us to determine whether inductive model discovery techniques have potential to identify new dynamic epidemiological models. Perhaps more importantly, this exercise could also tell us whether model discovery can enrich and nuance our understanding of existing study systems by uncovering a role for nonlinear mechanisms that were previously not considered. In the next section, we first apply SINDy to model rediscovery of a compartmental model with a seasonally varying transmission rate. Then we apply SINDy to model discovery from infectious disease case notification data for measles, rubella and chickenpox, and compare it to a standard compartmental epidemic model.

## Results

### Model Rediscovery from Simulated Data

For model rediscovery we use a discrete-time Susceptible-Infectious-Recovered (SIR) model accounting for demographic processes (birth and death) and seasonal variation in the transmission rate:

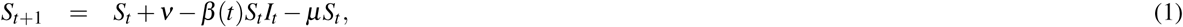

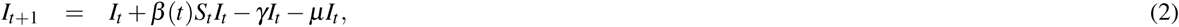

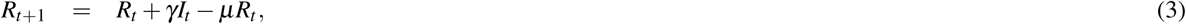

where *S*_*t*_ (*I*_*t*_, *R*_*t*_) is the number of susceptible (infectious, recovered) persons, *t* is the timestep, *ν* (*µ*) is the per capita birth (death) rate per timestep, *γ* is the per capita recovery rate per timestep, and *β* (*t*) is the seasonally-varying transmission rate with form given by

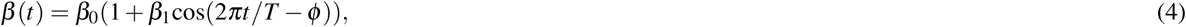

where *T* = 1 year is the period of the oscillation and *ϕ* is the phase shift corresponding with the seasonal behaviour of the transmission rate. We interpreted one timestep to correspond to one week. The mass action incidence term (*β*_*t*_*S*_*t*_*I*_*t*_) appears in both the *S*_*t*+1_ and *I*_*t*+1_ equations. Note that we may treat the state variable *R* as redundant since it does not appear in the other equations, and thus we may exclude it from simulations.

We used a discrete-time instead of a continuous-time (differential equation) model because SINDy necessitates evaluating the derivative of the input data, which for a continuous time model applied to empirical data can yield noisy and unpredictable values, making it difficult for the sparse regression algorithm to obtain a global minimum. In contrast, applying SINDy to a discrete-time modelling framework lends itself naturally to the weekly temporal resolution of the empirical data we used. Hence we use a discrete-time compartmental model for both model rediscovery as well as for model discovery from empirical data. However, we note that model rediscovery with SINDy also works with a continuous-time compartmental model.

We solved the discrete-time SIR model numerically and added Gaussian noise to the output state variables to generate a simulated dataset. SINDy was then applied to the simulated dataset using a function library consisting of all polynomials involving *S*_*t*_ and *I*_*t*_ up to and including second order, for both constant and seasonally-varying coefficients (see Methods). This process was repeated for a range of noise magnitudes. ‘Coefficient’ refers to the weighting assigned by SINDy to a given term, but we will use the terminology interchangeably.

For sufficiently small noise, SINDy was able to recover the original model with a very high accuracy by correctly recovering the coefficients of the SIR model (Figure 2). However, at higher (but still relatively small) noise magnitudes, SINDy begins to overfit the noise in the simulated data and, while it still correctly identifies the seasonally-varying mass action mixing term as having the largest coefficient, it also incorrectly identifies other nonlinear terms as having a role in the model, such as 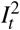 (Figure 2). The relatively small values for noise at which these spurious terms are introduced illustrates the challenges that noisy data present for SINDy. (However, it is also possible that other methods for regularization not explored here might enable SINDy to fit the data well even for the higher noise case.) Results for other noise levels appear in Supplementary Figures 1-4.

**Figure 2.**
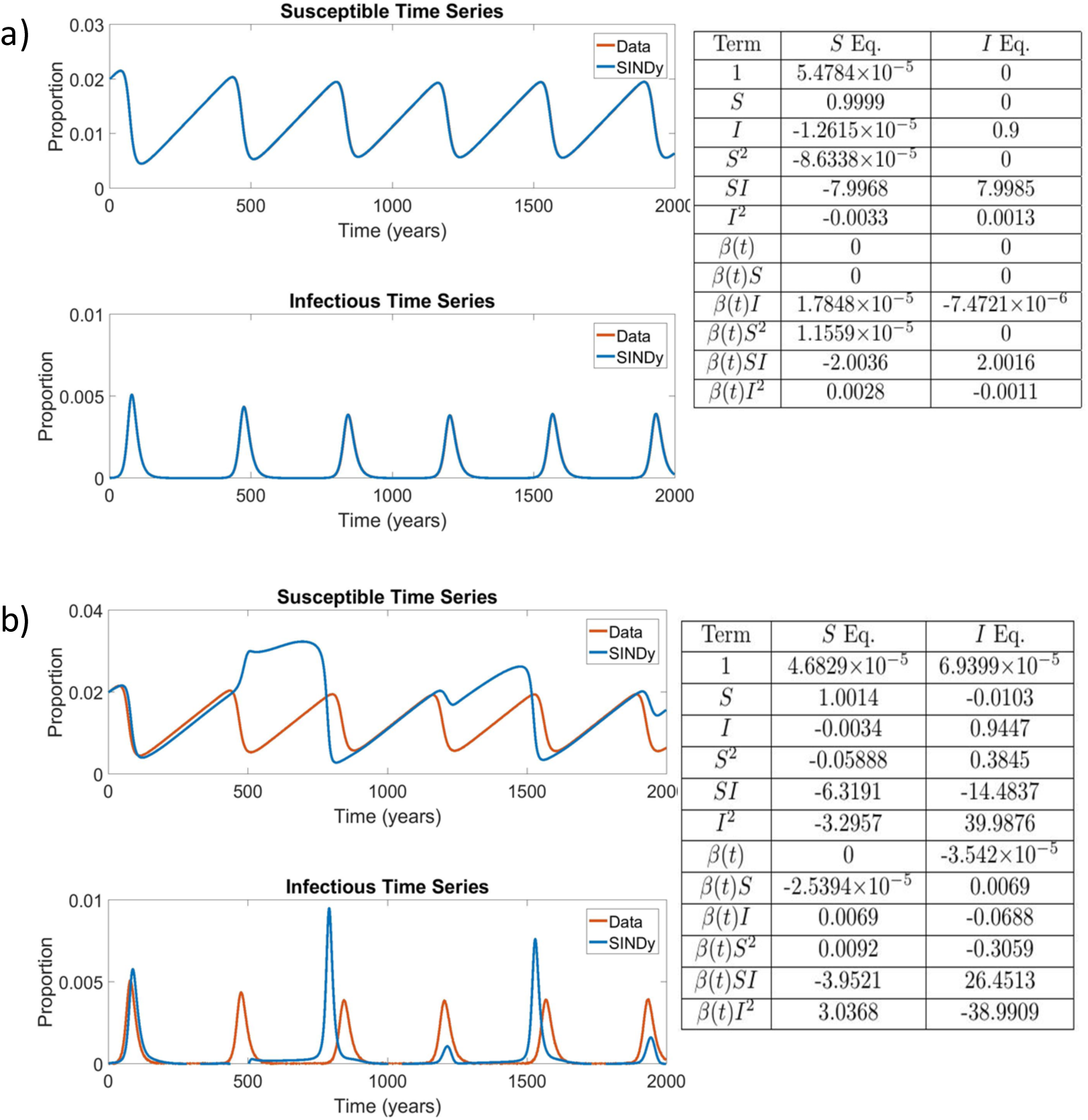
Comparison of the simulated SIR model with additive noise of *ε* = 1 × 10^−7^ (top) and *ε* = 2 × 10^−5^ (bottom) with the corresponding coefficients of terms in the discovered model. *β*_0_ = 8/wk, *β*_1_ = 0.25, *γ* = 0.1/wk, *µ* = *ν* = 5.4795 × 10^−5^/wk. The results in the table display the SINDy-discovered coefficients of the corresponding terms in (Eqs. 1 - 3). In panel (a) the Data and SINDy curves are very closely overlapping, hence the appearance of only a single blue curve.

### Model Discovery from Empirical Data

We studied three datasets consisting of case notifications for measles in England and Wales (1948-1967); chickenpox in Ontario, Canada (1946-1967); and rubella in Ontario, Canada (1946-60) (Figure 1). Each provides a different example of an attractor class: measles is biennial, chickenpox is predominantly annual, and rubella is multiennial with a weaker annual signatures as well^24,26^. Hence, these data represent an interesting test of whether a SINDy model discovered from a library of annually-forced transmission rates can generate a model that can predict not only annual but also biennial or multiennial dynamics. Our baseline analysis used a second-order polynomial library. However, we also tried a third-order polynomial library, which in principle allows capturing more features of the data but also risks more overfitting of noise. The data in Figure 1 was first smoothed to reduce the risk of overfitting (see Methods).

The datasets describe the case incidence (number of new cases per week), but our SINDy state variables concern the prevalence of epidemiological states: the number of susceptible individuals (*S*_*t*_) and infectious individuals (*I*_*t*_, the infection prevalence) at any given time *t*. Hence we converted case incidence to case prevalence to obtain the required input time series of the number of infectious individuals (see Methods).

We do not have data on the temporal dynamics of susceptible individuals, so we reconstructed a time series of the number of susceptible individuals using a standard method (see Methods)^37^. The method requires the initial number of susceptible persons, *S*_0_, and this initial condition can strongly influence disease dynamics. The number of susceptible individuals in a given year is not known for any of our historical datasets. Similarly, as mentioned in the Introduction, coefficients below a certain sparsity threshold value (*λ*, the “sparsity knob”) are removed from the library at each iteration of SINDy. But, there is no *a priori* knowledge to guide the selection of the value of *λ*. Hence, we applied SINDy to each point of the *S*_0_ − *λ* parameter grid. The Akaike Information Criterion (AIC) score of the model identified at each point on the parameter grid was computed to give us a way of measuring model parsimony across the grid^38^. To quantify sparsity of the coefficient matrix we introduced a *sparsity index* which is the ratio of the number of library functions with zero coefficients to the total number of functions in the library:

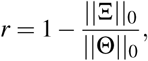

where Ξ is the set of coefficient vectors and Θ is the collection of library functions.

We found that SINDy discovered models that reflect the observed dynamics of infectious individuals for both measles and chickenpox (Figures 3 and 4). This includes both the biennial attractor of measles and the annual attractor of chickenpox. The models are not very sparse (*r* = 0.25) although the remaining coefficients differ greatly in the magnitudes that SINDy assigns to them. For measles, SINDy assigned the largest coefficients to the bilinear incidence terms *SI* and *βSI*, corresponding to constant and seasonally varying mass-action incidence terms respectively, as in the discrete-time SIR model (Figure 3).

**Figure 3.**
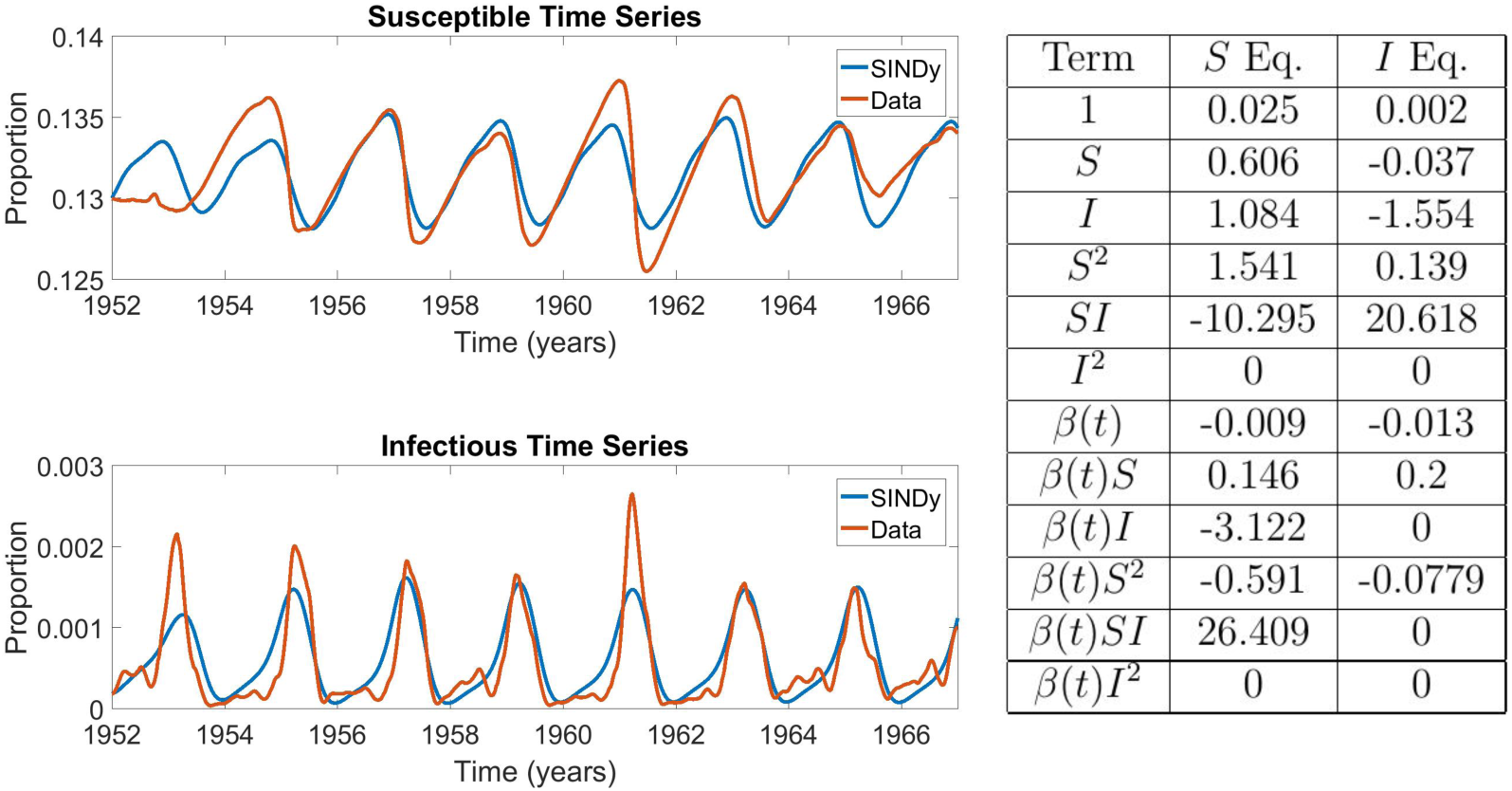
Comparison between measles incidence data and coefficients of the best SINDy-discovered model using a function library of polynomials up to 2nd order, and showing the model with the lowest AIC scores across the *S*_0_ − *λ* parameter grid. The discovered model accurately replicates the biennium present in the data in both the susceptible and infection classes. It also identifies a strong dependence on the *SI* and *βSI* cross terms, the driving terms behind the mass action incidence mechanism present in the SIR model. The sparse regression excluded six terms, giving *r* = 0.25. *S*_0_ = 0.11286 and *λ* = 0.00517. The parameter grids appear in SI Appendix, Figure 5. The results in the table display the SINDy-discovered coefficients of the corresponding terms in (Eqs. 1 - 3).

**Figure 4.**
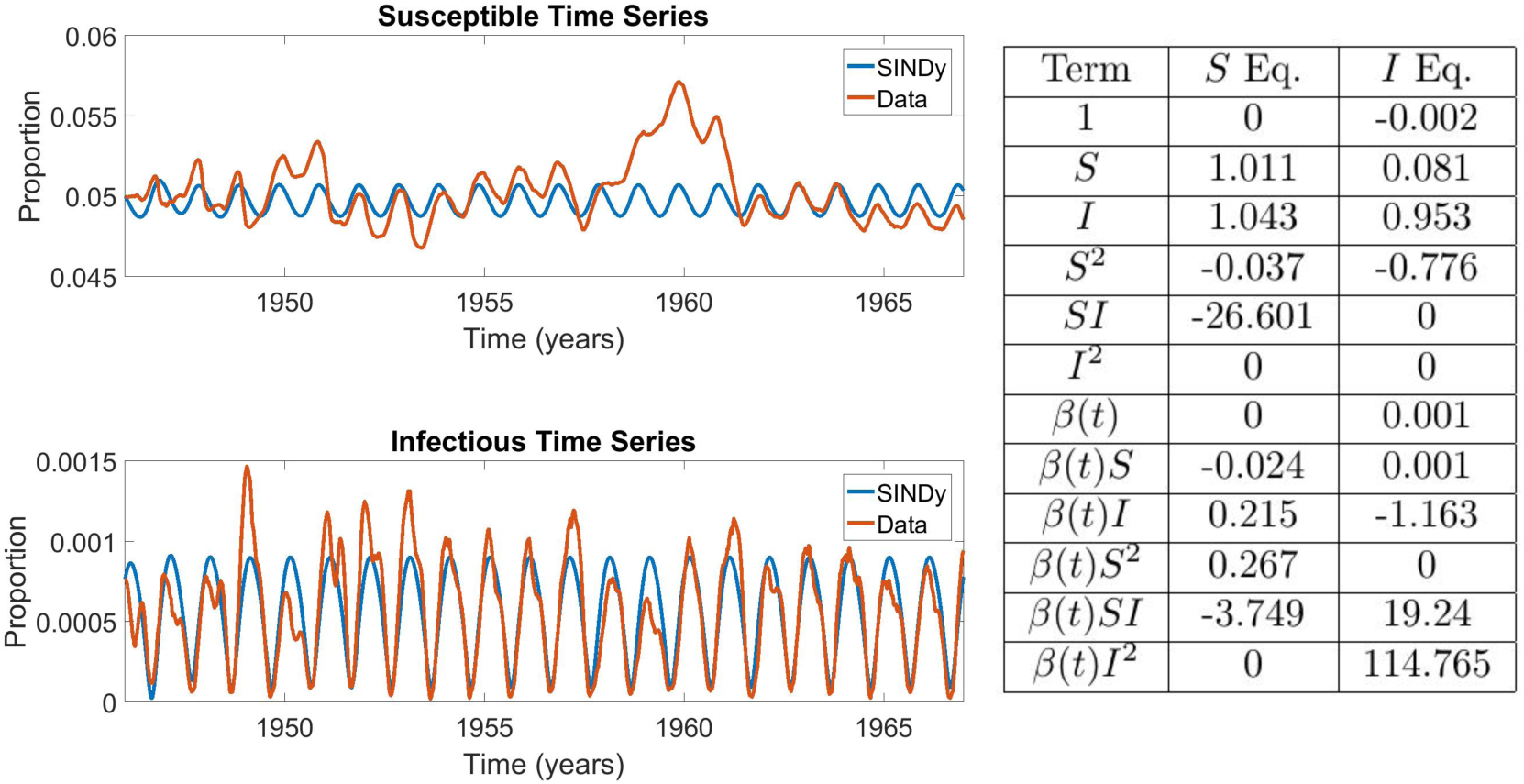
Comparison between chickenpox incidence data and coefficients of the best SINDy-discovered model using a function library of polynomials up to 2nd order, and showing the model with the lowest AIC scores across the *S*_0_ − *λ* parameter grid. The discovered model accurately replicates the annual cycle present in the data in both the susceptible and infection classes. As in the measles case, it also identifies a strong dependence on the mass action incidence term in both the *S* and *I* equations. Note also that the coefficient of *S* and *I* in their respective equations are close to 1, as expected in discrete disease models. The sparse regression excluded six terms, giving *r* = 0.25. The parameter grids appear in SI Appendix, Figure 7. The results in the table display the SINDy-discovered coefficients of the corresponding terms in (Eqs. 1 - 3).

SINDy also assigned large values for the *SI* and *βSI* coefficients for chickenpox, although the *β II* term was assigned the largest value overall, suggesting overfitting (Figure 4). The model that SINDy discovered for rubella was more sparse (*r* = 0.7), and SINDy also assigned the largest coefficients to the mass-action mixing terms *SI* and *βSI*, although the model predicts an annual attractor instead of the observed multiennial attractor (Figure 5). This occurs despite the fact that models with bilinear incidence are capable of generating multiennial attractors a rubella^24,26^. We reserve a more complete discussion of overfitting and the interpretation of SINDy model coefficients for a following subsection.

**Figure 5.**
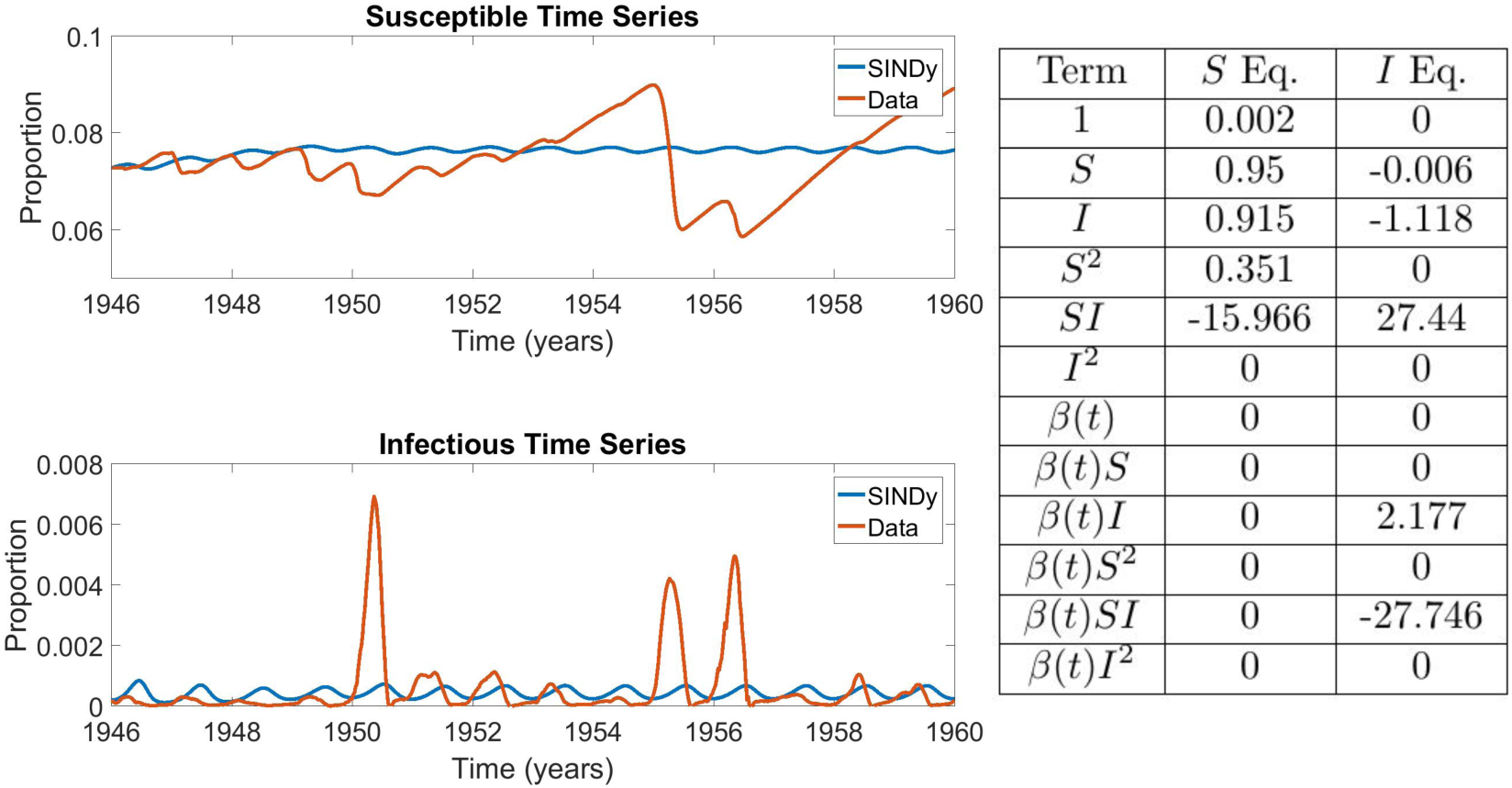
Comparison between rubella incidence data and coefficients of the best SINDy-discovered model using a function library of polynomials up to 2nd order, and showing the model with the lowest AIC scores across the *S*_0_ − *λ* parameter grid. The algorithm was unable to discover a model that exhibited the multi-annual cycle observed in the data, instead returning an annual cycle. Despite this, strong dependence on the mass action incidence term is again present. The sparse regression excluded 14 terms, giving *r* = 0.7. The parameter grids appear in SI Appendix, Figure 10. The results in the table display the SINDy-discovered coefficients of the corresponding terms in (Eqs. 1 - 3).

The fit between model and data for measles and chickenpox is low compared to many well-controlled examples from physics^10^, but it is comparable to the agreement achieved in many studies in epidemiological modelling^39^. This is because biological and social systems are complex and have multiple nonlinear feedbacks, in addition to numerous environmental heterogeneities. We discuss the case of rubella further in a following subsection on using power spectral densities.

The SINDy-discovered models confirm that seasonal forcing plays an important role in epidemics of childhood infectious diseases. The seasonally forced transmission rate (BSI) is always the first or second largest term in the discovered models. This provides a separate line of evidence in support of deductively derived models^24,26^ that support the role of seasonal variation in the transmission rate. Also, despite the fact that seasonal forcing occurs at a period of one year, SINDy can recover a seasonally forced model that exhibits a biennial attractor, hence these results provide another line of evidence that seasonal forcing can generate attractors of multiple different periods^24^, some of which are epidemiological relevant. The emergence of a biennial attractor from seasonal (annual) forcing happens due to the interplay between forcing dynamics, the gradual build-up of new susceptible individuals through births, and its rapid depletion during epidemic periods.

### Power spectral density

The results for rubella (Figure 5) highlight a limitation of using goodness-of-fit to time series as the criterion. As noted, a bilinear incidence term can generate multiennial attractors under seasonal forcing, but SINDy selected a model that generates an annual attractor under a second-order library. In terms of qualitative descriptions of the data, a model that generates multiennial attractors of the right frequency but with epidemic peaks at different years than what is observed in the dataset is perhaps more desirable, and says more about underlying epidemiological mechanisms, than a model that simply yields an annual attractor.

Hence, we developed a modification of our approach such that the goodness-of-fit between the power spectral density (PSD) of the model and the data is used as the criterion for assessing models, instead of fit between the prevalence time series (see Methods). Using this approach for rubella with a second-order polynomial library yields a model that reproduces the large multiennial outbreaks observe in the empirical dataset, as well as the notable tendency for seasonal rubella incidence to ramp up slowly in the first part of the season, but decline very quickly in the second part (Figure 6). The discovered model also includes a strong contribution from the mass-action incidence term *β* (*t*)*SI*.

**Figure 6.**
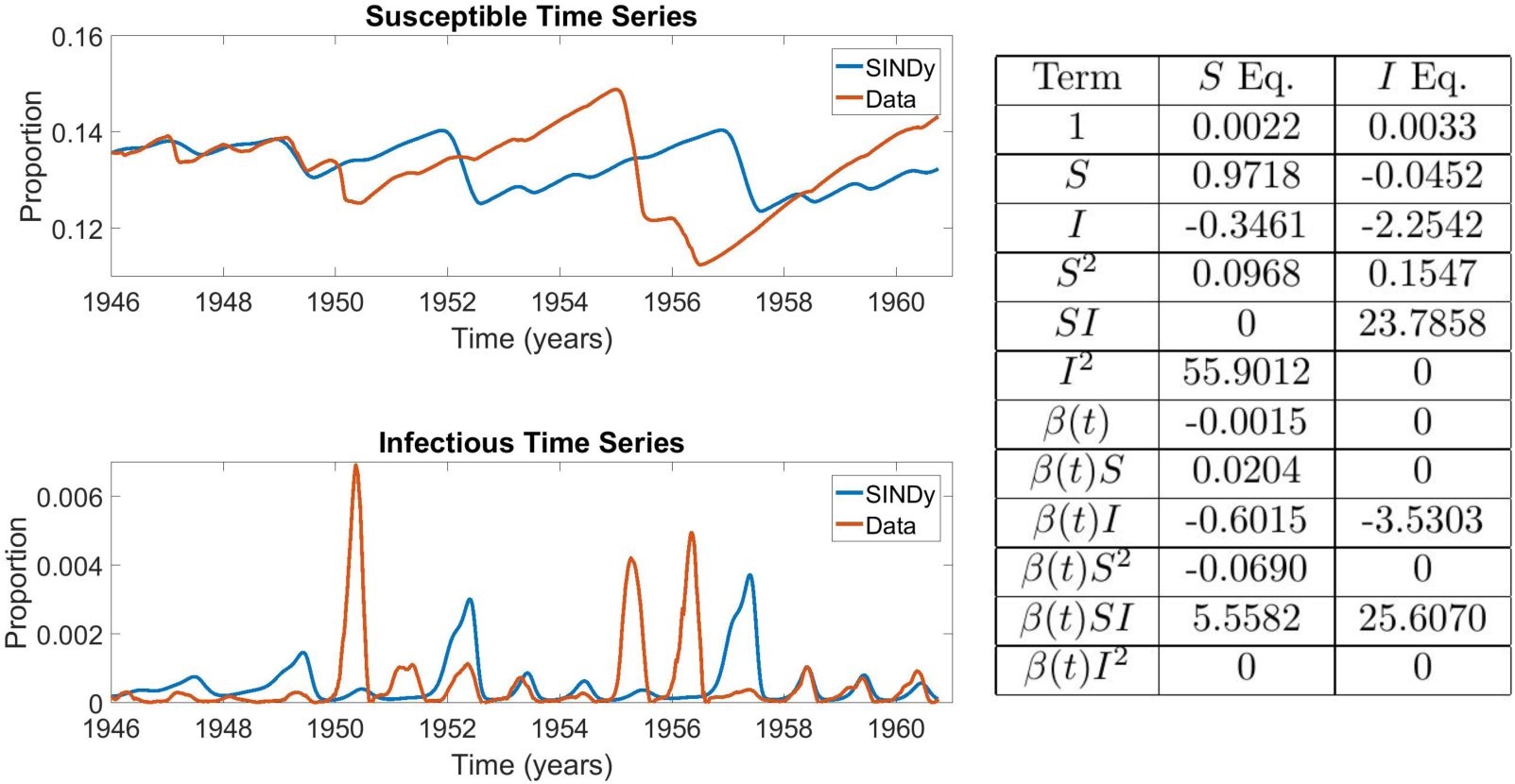
Comparison between rubella incidence data and coefficients of the best SINDy-discovered model using a function library of polynomials up to 2nd order with power spectral density as the criterion for AIC selection. The results in the table display the SINDy-discovered coefficients of the corresponding terms in (Eqs. 1 - 3).

The estimated power spectral densities for the empirical data and the best SINDy models appears in Supplementary Figures 5-7 for all three diseases. These figures show that dominant peak of the SINDy model PSD matches that of the empirical data, for all three diseases: 1 year for chickenpox, 2 years for measles, and 5 years for rubella. However, the SINDy model for chickenpox fails to capture some of the lower frequencies present in the empirical data.

### Interpretation of SINDy model coefficients

Inspection of the SINDy coefficients for the *I* variable in the baseline measles and chickenpox analyses (Figures 3, 4), as well as the PSD analysis for rubella (Figure 6) suggests that SINDy is producing meaningful models for the measles baseline and rubella PSD analyses, but could be overfitting the data in the chickenpox baseline analysis. (As noted above, SINDy exhibits a poor fit in the rubella baseline analysis, Figure 5.) We focus our discussion on the *I* equation for which observational data are available to SINDy, unlike the case for the *S* equation where SINDy must fit reconstructed data.

The SINDy models for the measles baseline and rubella PSD analyses exhibit biologically meaningful coefficients. We observe order 10^1^ two-variable terms pertaining to transmission (such as *SI*); order 10^0^ single-variable terms pertaining to demographic processes (such as *S, I*); and order 10^−1^ or 10^0^ *S*^2^ terms. All other terms are of order 10^−2^ or smaller. The relative orders of magnitude of the transmission and demographic-related terms are expected given that epidemic processes occur an order of magnitude faster than demographic processes. We also note that the SINDy models include both SI (constant) and BSI (seasonally varying) terms. This is also expected since the transmission rate in the SIR compartmental model *dI/dt* = (*b*_0_ + *b*_1_ sin(*t*))*SI* + … includes both seasonally varying and constant contributions to the total transmission rate^24,26^. Their relative magnitude varies depending on how important seasonal forcing is for dynamics. The sign of the BSI term may be positive or negative, depending on the phase shift.

The presence of the positive *S*^2^ term in the *I* equation both the baseline measles and rubella PSD models is notable. Adding a term of the form 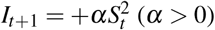 to the *I* equation has the effect of slowing down and linearizing the epidemic curve in the early stages of the epidemic. This is because the transmission portion of the equation changes from *I*_*t*+1_ = *βS*_*t*_*I*_*t*_ + … (as in the SIR model) to 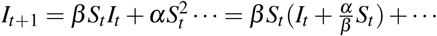. As a result, the force of infection changes from *β I*_*t*_ to 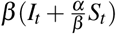. In the early stages of the outbreak *S*_*t*_ ≫ *I*_*t*_ and *S*_*t*_ ≈ *S*_*cst*_ is approximately constant, hence, the force of infection is approximated by 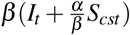; this functional form has a smaller slope with respect to changes in *I*_*t*_, meaning that the epidemic will growth subexponentially. We also note that *α <* 0 has the opposite effect and could even make incidence of new infections decline, if *α* is large enough.

The *S*^2^ coefficient is positive in the rubella PSD and measles baseline SINDy models, but absent for the rubella baseline model and has the wrong sign for the chickenpox baseline model. The empirical case notification data for measles and rubella show a corresponding pattern of a slower initial growth (in the case of measles, driven partially by small outbreaks in even years), followed by a more rapid decline after the peak of the outbreak. The SINDy model time series for the baseline measles and rubella PSD analyses share this feature as well. However, this pattern is not apparent in the chickenpox data or in the baseline chickenpox SINDy dynamics. Moreover, in contrast to the measles baseline and rubella PSD SINDy models, conventional compartmental models always show a symmetric epidemic curve, or faster growth followed by a slower decay. This demonstrates an advantage of the SINDy models over classic compartmental models for capturing subexponential epidemic growth in the early stages of an epidemic–a time when accurate projection of new cases is most important. It also suggests a simple way that early subexponential growth could be captured phenomenologically in simple compartmental models without the need to resort to more complicated models such as spatial models (which can also capture subexponential growth^40^,^41^).

The *I* equation for the baseline chickenpox SINDy model is strongly dominated by the seasonally varying *BII* coefficient. This suggests that SINDy is overfitting the chickenpox data. This outcome is made more likely by the fact that the chickenpox data exhibit a simple annual cycle that can be better explained by a sinusoidal oscillation than a nonlinear transmission model. In comparison, measles dynamics show a two-year periodicity, and rubella dynamics show a multi-year periodicity. In both latter cases, interaction between nonlinear mass-action infection dynamics, vital dynamics, and seasonal forcing appears necessary to explain their rich dynamics^24^,^26,27^.

### Effect of changing sparsity knob *λ*

Our approach applies SINDy to a grid of possible values for the initial number of susceptible individuals *S*_0_ and the sparsity knob *λ* which determines the threshold for removing terms from the library in each iteration of SINDy. The sparsity knob is particularly important because if it value is set too low, the discovered model will include most (or all) of the functions in the library, resulting in overfitting. Conversely, if the threshold is set too high then features required to emulate the dynamics of the system may be removed, resulting in a model that does not resemble the data in a meaningful way.

Hence, in order to balance sparsity with goodness-of-fit, we focussed on the models that yielded the lowest AIC score across the grid for the foregoing results (Figures 3-5). This approach shows there is an optimal region in the *S*_0_ − *λ* plane that ensures the SINDy algorithm can generate regularized, accurate models from empirical disease data (Supplementary Figures 8-15). For instance, in the case of measles, the best AIC corresponded to a model using an initial susceptible value of *S*_0_ = 0.11286 and a sparsity knob of *λ* = 0.00517 (Figure 3). In contrast, a much lower sparsity setting (*λ* = 0.0001) discovers a model that is overfitted to apparently random features of the data and that includes all the terms of the library, whereas a much higher sparsity setting (*λ* = 0.1) discovers a model that exhibits annual attractor instead of the characteristics biennial attractor of measles (Supplementary Figure 9).

### Third-order polynomial library

When a third-order library is used instead, the discovered models for measles and chickenpox capture the observed epidemiological dynamics as well as the second-order library does, and the models have similar sparsity indices (Supplementary Figures 16-18). SINDy continues to assign significant weight to the bilinear incidence terms *SI* and *βSI*, but SINDy assigned even stronger weight to the *S*^2^*I* and *SI*^2^ terms (and their corresponding seasonal terms). This could indicate overfitting, or it may indicate a more general nonlinear incidence mechanism in the underlying system. It has been shown that an incidence function of the form *S*^*p*^*I*^*q*^ (where *p, q >* 0) may more adequately represent some endemic cycles, a form that is present when a 3rd order polynomial library is used^42^,^43^. In the case of rubella, SINDy generates a model that captures the multiennial attractor observed in rubella dynamics (Supplementary Figure 15) although the model is not very sparse (*r* = 0.15) and is strongly dependent on trilinear incidence terms *S*^2^*I* and *I*^3^. Parameter planes for *S*_0_ and *λ* and examples of predictions for very low and high sparsity thresholds appear in Supplementary Figures 19-27.

### Comparison with compartmental epidemic models

We compared the models discovered by SINDy to the classical discrete-time SIR compartmental model with seasonal forcing and mass-action mixing. We fitted Equations (1)-(4) to the case prevalence and reconstructed susceptible time series for all three infections by sweeping across parameter grids and minimizing the least-squared error between model and data time series (see Methods). We thereby inferred the parameter values *ν, µ, β*_0_, *β*_1_ and *γ* and compared these parameter values to the coefficients of functions determined from SINDy for the models in Figures 3-5.

These comparisons show that SINDy tends to select the coefficients corresponding to mass-action mixing, often with seasonal forcing as well, and that these coefficients have a similar magnitude and sign as the inferred parameters of the compartmental SIR model (Figure 7). The similarities are strongest for measles and chickenpox. This demonstrates that SINDy is effective in capturing the theoretical principles of mass action incidence–a core mechanism of most epidemiological models–as well as seasonal forcing, which is required to explain endemic patterns of many childhood infectious diseases in the pre-vaccine era. The SINDy models also depend on the linear terms for each of the *S* and *I* equations. Usually these have a similar magnitude and sign as in the SIR model. These terms correspond to vital dynamics (births and deaths). However, several other features are noticeably different from the inferred SIR model. For instance, SINDy often infers a different magnitude and/or sign than the SIR model, particularly for the baseline rubella analysis that provides a poor fit to the empirical case notification data (Figure 5).

**Figure 7.**
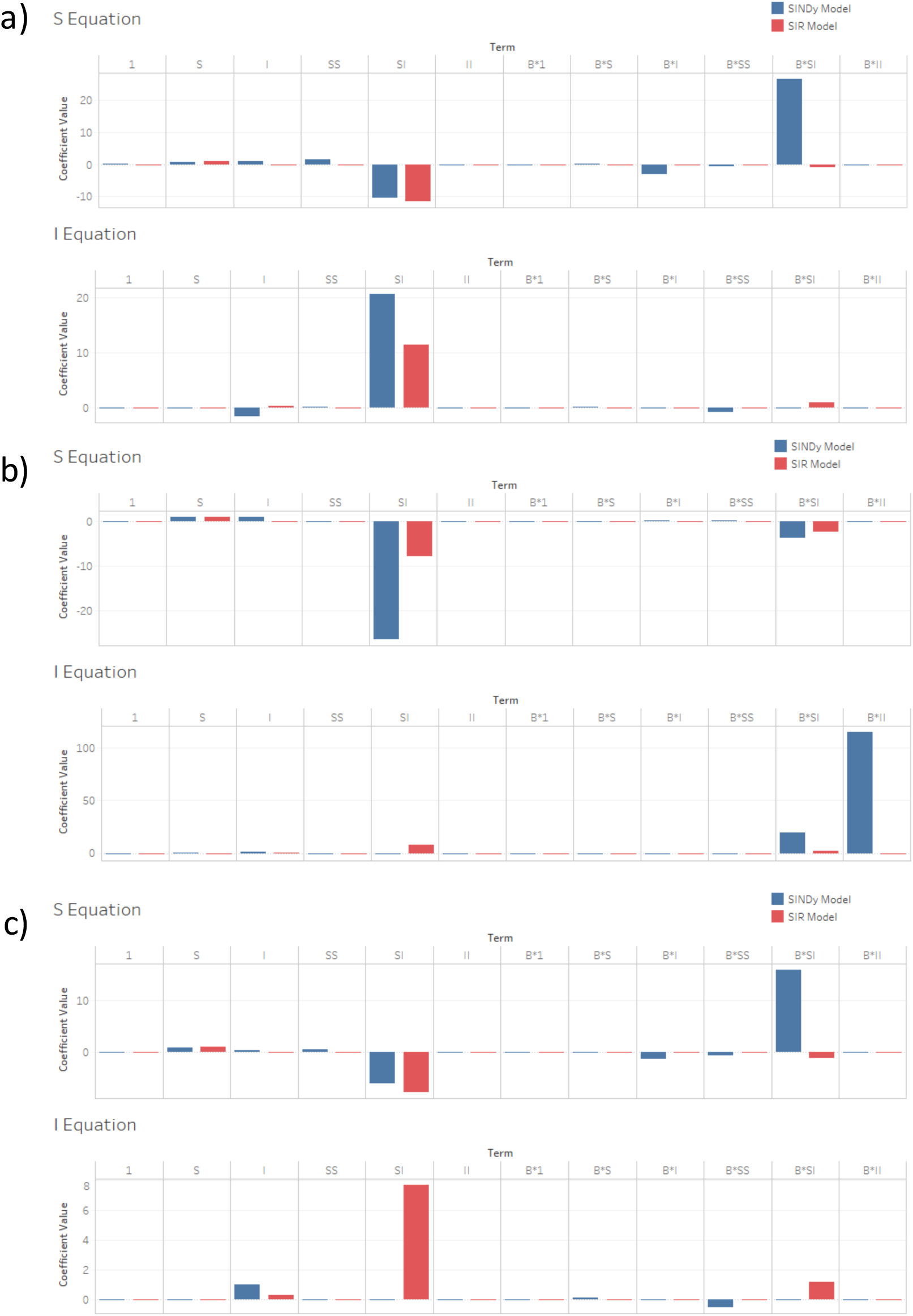
Comparison of coefficients between SINDy-discovered model (using a function library of 1st and 2nd order polynomials) and fitted SIR model for (a) measles, (b) chickenpox and (c) rubella.

### SINDy model can predict qualitative shifts in empirical measles dynamics out-of-sample

To test whether models discovered by SINDy can predict real-world dynamics, we studied the ability of the second-order SINDy baseline measles model (Figure 3) to make out-of-sample prediction of regime shifts in measles dynamics. We used a classic example of nonlinear measles dynamics in the United Kingdom^24^,^26^. During 1948-1967, the recruitment rate of new susceptible individuals in England & Wales was roughly constant and measles incidence exhibited a clear biennial pattern (Figure 1). However, the recruitment rate of new susceptible individuals dropped significantly in 1967, due to the introduction of mass vaccination and a decline in birth rates, causing dynamics to shift suddenly to a more irregular pattern dominating the time period 1968 to 1988^24^,^26^. Previous research has shown that simple compartmental epidemic models can predict this shift, as well as the patterns observed before and after the shift^24^,^26^. These models characterize the dynamics from 1948-1967 as a biennial attractor^24^ that is relatively stable to perturbations from noise (on account of the fact that the period of the biennium’s Floquet multiplier–which predicts response to noise–is two years and thus exactly matches the two-year period of the biennial attractor^26^. The dynamics from 1968-1988 are characterized as an annual attractor that is more easily perturbed by noise (on account of the Floquet multiplier of the annual attractor having a non-annual period greater than one year^26^). These differences have implications for the power spectra of the time series of infection incidence. In the power spectra of both the classical models and the empirical data, we observe a shift from a dominant peak at two years and very little power elsewhere in the spectrum (except for a supporting annual peak) during the biennial era, to a dominant peak at one year and a second peak at a period of approximately 2.5 years (corresponding to the period of the attractor’s Floquet multiplier), during the era of irregular dynamics^26^.

The second-order SINDy model for measles was discovered from the incidence patterns observed during the biennial era (Figure 3). We hypothesized that if the SINDy model is discovering real-world mechanisms and not over-fitting the data, it should predict the same transition observed in the empirical data and predicted by previous models^24^,^26^. To test this hypothesis, we reduced the susceptible recruitment rate in the SINDy model (the coefficient of the *S* term) to match the drop in the susceptible recruitment rate observed in the United Kingdom from 1948-1967 to 1968-1988 and we compared the time series and power spectra of infection incidence and susceptible individuals before and after the drop. We also added white noise to the simulations to test the response of the attractors to noise. We found that the SINDy model predicts the same transition. In the biennial era, the SINDy model predicts a stable biennial cycle that is relatively robust to noise: the time series of infection incidence shows a clear biennial pattern and the power spectrum shows a strong peak at a period of two years, a supporting peak at one year, and little else (Figure 8 and 3). In simulations where the coefficient of the *S* term is reduced to capture a drop in the recruitment rate, the SINDy model predicts a noisy annual cycle: the time series of infection incidence shows a dominant annual signature in the presence of significant irregularities and perhaps an envelope of a non-seasonal periodicity, and the corresponding power spectrum shows a strong annual peak with a secondary peak at a higher period, caused by the response of the annual attractor to noise. The time series also resemble the empirical post-vaccine dynamics^24^,^26^. Hence, the SINDy model is predicting the empirically observed regime shift in measles dynamics out-of-sample. (Predicted susceptible dynamics are also shown in Figures 8, 9 but were not analyzed in Refs.^24^,^26^. Ten additional stochastic realizations with different starting random number seeds and their power spectra appear in the SI appendix: Figures 28-37. These show similar patterns to Figures 8 and 9.)

**Figure 8.**
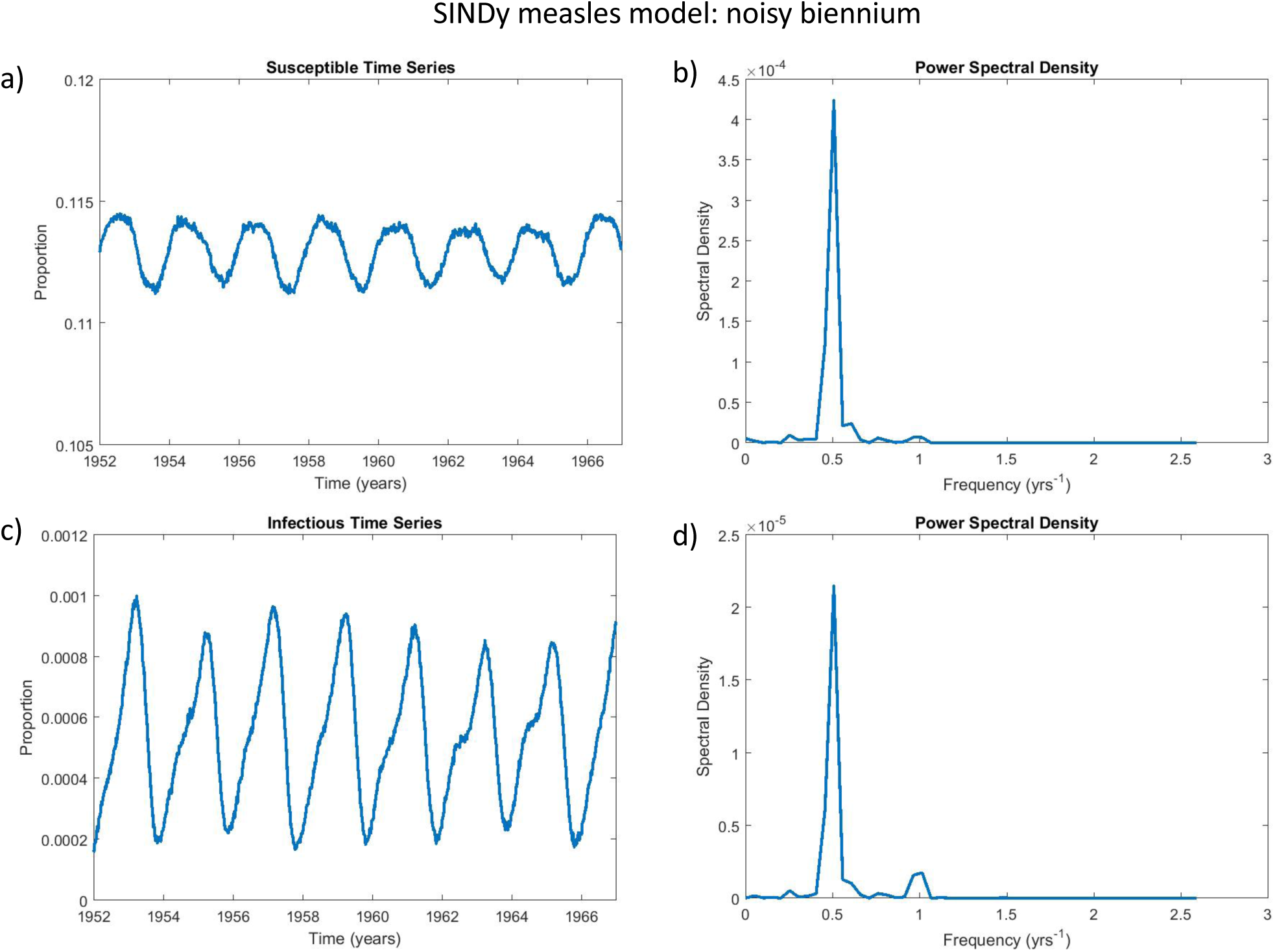
The SINDy measles biennium is robust to the addition of noise. This figure depicts the simulated timeseries of the SINDy measles model under additive noise: subpanels show the proportion of susceptible (a) and infected (c) individuals over time and the corresponding power spectral density plots for the susceptible (b) and infectious (d) time series. The power spectral density plots show strong power at a frequency of 0.5*/*year and a lesser peak at 1*/*year, corresponding to a prominent biennial cycle. White noise with a coefficient of 1.5 × 10^−3^ was added to the right-hand side of the SINDy-discovered system of differential equations to generate these plots. See Methods for details about computation of the power spectral density.

**Figure 9.**
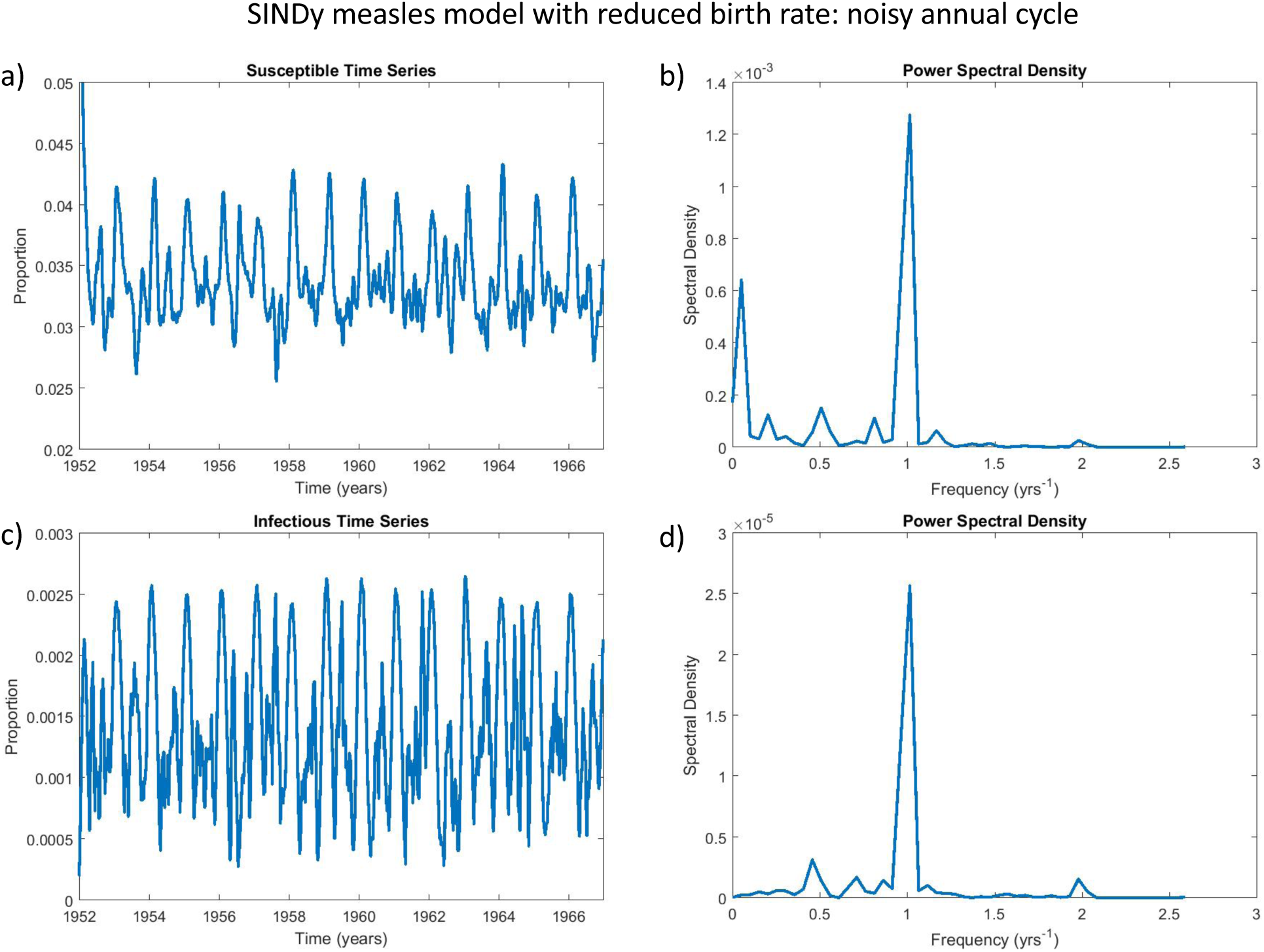
The SINDy measles model predicts a noisy annual attractor when the recruitment rate is reduced. This figure depicts the simulated timeseries of the SINDy measles model with a reduced birth rate, under additive noise: subpanels show the proportion of susceptible (a) and infected (c) individuals over time and the corresponding power spectral density plots for the susceptible (b) and infectious (d) time series. The power spectral density plots show strong power at a frequency of 1*/*year corresponding to a prominent annual cycle, as well as power at lower frequencies corresponding to multiennial cycles^26^. White noise with a coefficient of 1.5 × 10^−3^ was added to the right-hand side of the SINDy-discovered system of differential equations to generate these plots. See Methods for details about computation of the power spectral density. To simulate a reduced recruitment rate of newly-born susceptible individuals from 2.60/year to 1.36/year (expressed as the total fertility rate of susceptible offspring) between 1948-1967 and 1968-1988 in the United Kingdom due to falling birth rates and mass vaccination^44^ the coefficient of *S* was changed from 0.606 (Figure 3) to 0.317.

## Discussion

Model discovery generates models inductively from data, using minimal prior knowledge about the system. This differs from the deductive approach that currently dominates model development in most fields, including theoretical epidemiology. Here we demonstrated that Sparse Identification of Nonlinear Dynamics (SINDy) can discover dynamical system models from empirical data on childhood infectious diseases from the pre-vaccine era. These inductively derived models (1) reproduce the observed dynamics of measles, chickenpox and rubella, (2) recover prominent features of deductively derived models, such as the fundamental mechanisms of mass-action mixing and seasonal forcing that underpin decades of research in theoretical epidemiology, (3) confirm the important role of seasonal variation in the transmission rate for dynamics of childhood infectious diseases, and (4) can predict regime shifts in dynamical patterns for measles, out-of-sample. Hence, our results show that model discovery methods could lend insight to both model creation as well as understanding of epidemiological mechanisms. Because this approach to developing the dynamic models is fundamentally different from the conventional approach, it raises the possibility that real-world mechanisms could be discovered that would otherwise be difficult or impossible to find through conventional ‘deductive’ modelling.

However, we also note that the model discovered by SINDy appeared to be overfitted in the case of chickenpox. Also, we were not able to obtain a good fit to the empirical rubella time series data; this prompted us to fit the power spectral density instead, which yielded a reasonable qualitative agreement between model and data. The approach of using power spectral density as a fitting criterion might be desirable for describing the qualitative outbreak patterns of infectious diseases like rubella, which in our dataset exhibits large epidemics every 6-7 years interspersed with small annual epidemics. These aspects of our findings emphasize the challenges in applying SINDy to complex, noisy biological systems.

Limitations to the approach stem from data quality and availability, regression algorithms and criterion for selecting parameters like the sparsity threshold. Case notifications are typically under-reported even when they are available, and our approach requires reconstruction of the susceptible time series, which necessitates making assumptions^37^,^45^. SINDy may be sensitive to the method used to reconstruct the susceptible time series, and these methods might also bias SINDy toward selecting certain terms. Unfortunately, susceptible reconstruction is necessary due to the lack of high-quality longitudinal serological data, but a careful sensitivity analysis in future work could help us better understand the impact of differing methods of susceptible reconstruction.

Applying SINDy to vaccine era data will require further thought because the vaccination in the 20th century changed the dynamics of childhood infectious disease dramatically^24^,^26,45,46^ and introduced the additional dimension of human behaviour through vaccine decision-making^47^–^49^. Fortunately, data are increasingly abundant in an era of digital data, open sharing, and online social media^50^,^51^, although this does not necessarily translate into higher quality data. And, the risk of overfitting to noise is always present and will only increase as more data becomes available^52^. In this case, we tested whether the model was over-fitted by analysing its out-of-sample prediction of empirical measles dynamics. However, over-fitting could also be tested varying the window of the moving average filter or subsampling the data before filtering.

The sensitivity of SINDy to noise (despite improvements over previous algorithms in this respect) is another limitation, and was illustrated in our model rediscovery subsection. Re-discovered models may describe the datasets very well in the presence of noise, but the spurious inclusion of terms that do not appear in the original model suggests caution when interpreting models discovered from noisy empirical data. We hypothesize that noise is also the cause of unexpected terms in our models discovered from empirical data, especially for the results using a third-order library. Part of the solution to this problem will be improved regression approaches that are less likely to overfit noise. However, the other part of the solution is inevitably the thoughtful application of subject expertise by humans when interpreting discovered models.

Another limitation is the choice of functional basis. This might be the largest limitation, since human bias is introduced when formulating the function library that SINDy starts with^10^. Our choice of seasonally varying transmission rates and polynomial functions may be limiting the discovery of a more parsimonious and interpretable model based on other terms, at least in principle. The state variables themselves are also pre-defined and may not be the best choice to recover the dynamics of the data.

There remains much opportunity to develop further techniques that assist in applying sparse identification methods to epidemiological data. A more exhaustive literature review of current transmission modelling practices would aid in determining a functional basis that could successfully capture a sparse model using subset selection methods. There exist many modern adaptations on compartmental modelling of infectious diseases^53^,^54^ which incorporate functions that extend beyond a simple polynomial basis constructed from state variables. Specifically, a general nonlinear incidence (a transmission function of the form *S*^*p*^*I*^*q*^, where *p, q >* 0) could be explored. Extending the function library to include non-integer values for *p* and *q* may allow a more accurate representation of the transmission mechanism and result in a more parsimonious discovered model. In addition, the choice of a sinusoidal function for the transmission rate may not be optimal^24^. While an alternative based on forcing from the school year calendar was tested in our research, further analysis in this area is necessary.

In conclusion, we have shown that SINDy can be applied to epidemiological data to yield models that describe observed epidemic patterns and correspond well with canonical models like compartmental epidemic models. In the case of measles, it can also predict dynamical shifts out-of-sample. Although inductive model discovery methods come with their own set of challenges and limitations, their radically different approach to model generation suggests they can form a powerful complement to traditional modelling approaches, thereby improving our scientific understanding for many natural systems.

## Material and Methods

### Sparse Identification of Nonlinear Dynamics (SINDy)

This work builds on the sparse regression methods outlined in Ref.^10^. Given the recent advances in both compressed sensing^55^–^57^ and sparse regression^7^,^58^ it has become computationally feasible to extract system dynamics from large, multimodal datasets. These techniques rely heavily on the fact that many dynamical systems can be represented by governing equations that are sparse in the space of all possible functions. In this work we focus on dynamical systems that are given by a system of ordinary differential equations of the form

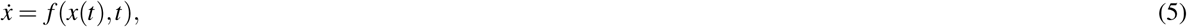

where *x*(*t*) = (*x*_1_(*t*), *x*_2_(*t*), …, *x*_*n*_(*t*)) represents the state of the n-dimensional system at time *t*, and *f* = (*f*_1_, *f*_2_, …, *f*_*n*_) is the sparse set of functions that dictate the dynamics of the system.

It is assumed that the time series data is sampled at points *t*_1_, *t*_2_, …, *t*_*m*_ for both *x* and 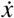, usually given as either data from simulations or empirical data from measurements. Depending on the system in question, numerical differentiation methods to approximate 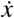 that are well-suited for the level of noise must be used. The method used in Ref.^10^ is total variation regularization^59^,^60^ that works well on a noisy system when only the state variables are available. Alternatively, a discrete adaptation of SINDy may be used, where the response of the system *f* (*x*_*t*_, *t*) is *x*_*t*+1_. Regardless, the time series data of the state variables and the response are represented by the matrices

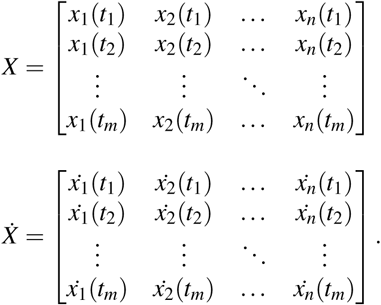

We then construct a library of linear and nonlinear candidate functions for the model, given prior knowledge of the system we wish to describe. Common choices for these functions are polynomial and trigonometric functions of the state variables, though other functions (e.g. exponential, rational) functions may be included as well. This function library is then evaluated at each time-step, generating the *m*× *p* matrix

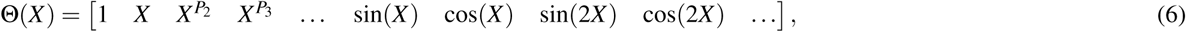

where 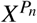 represents all possible polynomials of degree *n* that can be constructed by the state variables. Now, relying on the assumption that the derivative 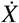 can be described by relatively few of the nonlinearities active in Θ(*X*), we may set up the sparse regression problem

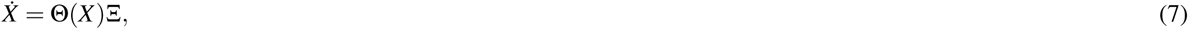

where Ξ = (*ξ*_1_, *ξ*_2_, …, *ξ*_*p*_) is a set of sparse coefficient vectors.

There are several current methods that have been developed to perform sparse regression. A common choice is the LASSO (least absolute and shrinkage operator)^7^,^58^, a regression method that promotes sparsity by applying an *l*_1_ penalty on the norm of the coefficient vector. However, this method does not scale well to large datasets. This paper utilizes an iterative method developed by Brunton et. al., as described below:

1. Perform a least-squares regression on the relation in Eq. [7].
2. Set all terms in Ξ that are less (in absolute value) than some threshold *λ* to zero.
3. Create new library Θ^*′*^, dropping functions that correspond to zero entries in Ξ.
4. Repeat steps 1-3 until equilibrium (i.e. no terms in Ξ are smaller in magnitude than *λ*), or some other stopping criteria is reached.

This yields the set of sparse vectors that provides an approximate solution to Eq. [7]. We can then reconstruct the *k*th row of the dynamical system by taking

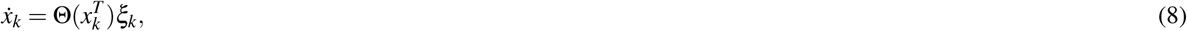

where 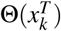 is the symbolic representations of the elements of *x*.

Finally, combining all of the rows of the discovered dynamical system results in the system of equations

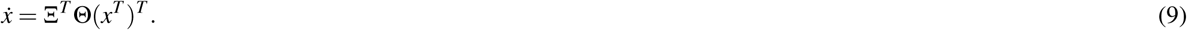

The code for this algorithm, along with several examples that demonstrate its application, can be found at Ref.^61^. The modified repository used for all computation done can be found at Ref.^62^.

### Applying SINDy to Epidemiological Systems

The application of data-driven model discovery methods to epidemiological systems presents a unique set of challenges. Firstly, incidence data is often subjected to noise at several levels, notably inconsistent reporting of infection cases^45^,^63,64^. In addition, the derivative data must be approximated using numerical methods, leading to another source of inaccuracy. Secondly, most compartmental epidemic models depend on the densities of both infected and susceptible individuals. However, temporal data of the seropositive individuals in a population would require extensive surveying and is frequency not available. Instead, several methods for reconstructing the susceptible class from the given incidence data are used (see subsection on Susceptible Reconstruction. When using up to second order polynomials, the function library we used was

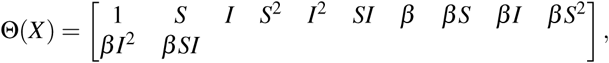

and when using up to third order polynomials, the function library used was

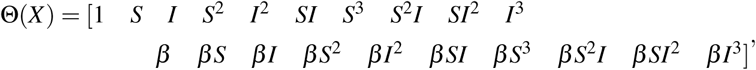

where *β* is the seasonally-varying transmission rate given in Eq. 22. When the model coefficients are given in figures describing SINDy-discovered models, this parameter is represented by *B*.

### Model Selection

There exists a group of statistical metrics that are used to balance goodness-of-fit with model complexity, called *information criteria*. These metrics are useful in the comparison and selection of models when first given a space of candidate models from which to choose. In the context of symbolic modelling this space is usually constructed from a functional basis, often heuristically defined given contextual theory^65–67^. Given a computationally tractable basis, each possible model would be fitted and the information criterion would be computed and used to select the model that best balances parsimony and predictive power. The information criterion used in this paper is called the Akaike information criterion (AIC)^38^ and is derived from use of maximum likelihood. The AIC value for a given candidate model *i* is defined by

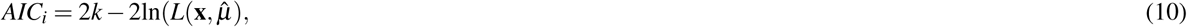

where *L* is the conditional probability of the observations **x** given the set of best-fit model parameters 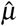, and *k* is the number of free parameters in the model.

### Data Sources and Preprocessing

Temporal data of infection incidence of various infections and time periods has been made available by numerous sources, often from governmental reporting programs. The three infectious diseases and the corresponding locations and time periods used for this study are measles in England and Wales from 1952-1967 (from Ref.^68^), chickenpox in Ontario (Canada) from 1946-1967, and rubella in Ontario from 1946-1960 (both from^26^). These diseases and time periods were chosen as they exhibit contrasting dynamic behaviour, most notably in the period of the epidemic cycle. For each of these diseases, the time frame chosen is before the vaccines for the respective diseases became commonly available. The raw data were also smoothed using a Savitzky-Golay filter^69^ of order 3 with a window length of 19 in order to reduce the risk of SINDy overfitting the data, resulting in the smoother time series shown in Figs. 2-6. Once this data was imported and both the time and case vectors were labelled, both the birth and population data (taken from^68^,^70–73^) were imported and linearly interpolated to be given per week, the same scale as the case notification data. Time series of birth rates for Ontario and the United Kingdom appear in Supplementary Figure 38.

### Discrete Time Model

In the limit as Δ*→ t* 0 the discrete model (Eqs. 1 - 3) converges to the continuous-time compartmental SIR model^26^,^30^. Applying SINDy to discover a continuous-time model involves determining the derivative vector 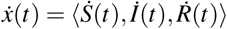. As this requires numerical differentiation of a potentially noisy system, valuable information can be lost. However, when using the discrete system, the response vector is *x*_*t*+1_ = ⟨*S*_*t*+1_, *I*_*t*+1_, *R*_*t*+1_ ⟩, which is simply the next data point and thus is implicitly available without numerical approximations. Hence, we used the discrete-time SIR model for our analysis. Solving Eq. 4 for *β* and iterating over empirical data for measles, chickenpox and rubella give the time series found in Supplementary Figure 25. Given that each of these diseases is most common among school-aged children^45^,^46,74^ it is unsurprising that the period corresponding with the lowest transmission is in the summer, followed by a peak in September correlating with a return to school. These findings are further discussed and confirmed by Refs.^74^,^75^, though more analysis by Ref.^45^ indicate the peak in transmission rate occurs several weeks earlier, alleging this effect may also be attributed to weather fluctuations.

### Susceptible Reconstruction

The simplest method for the reconstruction of the susceptible class is to iterate the equation

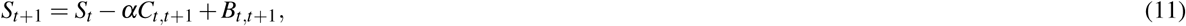

where *S*_*t*_ represents the number of susceptibles at the start of week *t, C*_*t,t*+1_ and *B*_*t,t*+1_ are the number of new cases and births respectively in week *t*, and *α* is the rate at which cases are reported (i.e. *α*^−1^ is the average proportion of all cases that are reported to the data collection agency)^45^. The idea behind this method is simple: each week the suceptible class grows by the number of new births into the population (in the absence of vaccination), and shrinks by the number of new infections. If the reporting rate *α* was well known, this relation would provide a good approximation. However, reporting varies significantly for different diseases and locations^64^ as well as changing temporally^37^. It is also difficult to estimate explicitly, due to the lack of serological data available.

An extension of this method is derived in Ref.^37^. They assume the discrete relation

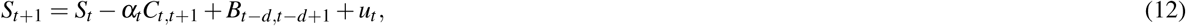

where *u* describes the additive noise 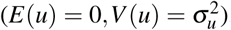, and *d* represents a short delay to allow for the period of time between birth and susceptibility to the disease. Now let *Z*_*t*_ describe the deviation from the mean 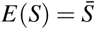 at week *t*, i.e.

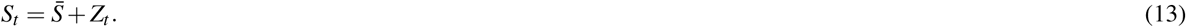

By substituting Eq. [13] into Eq. [12] we see that *Z*_*t*_ also satisfies the relation

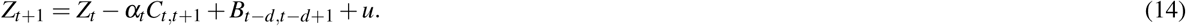

Iterating this expression results in the relation

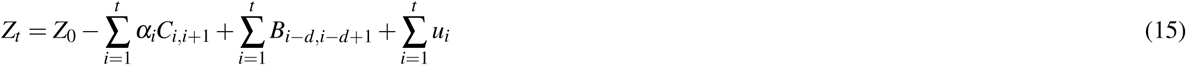

Ref.^37^ uses the simplifying notation

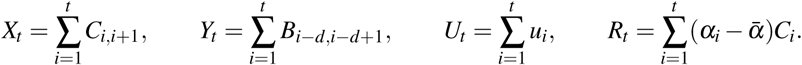

This simplifies Eq. [15] to

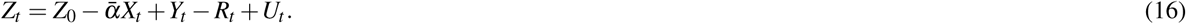

If it is assumed that the reporting rate is constant (*R*_*t*_ ≈ 0) and noise is negligible (*U*_*t*_ ≈ 0), this reduces to the linear relationship

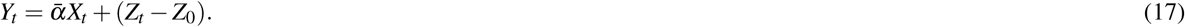

Hence, applying a linear regression to the cumulative births (*Y*_*t*_) against the cumulative cases (*X*_*t*_) provides an estimate for the residuals *Z*_*t*_ − *Z*_0_ and the average reporting rate 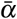.

We call this the ‘global regression method’. Applying this reconstruction method yields time series of of the proportion of susceptible individuals for the data in Figure 1 (Supplementary Figure 39). From these figures it can be seen that each reconstruction (especially for the chickenpox and rubella case notification data) suffers from local shifts in the mean, caused by the assumption that the reporting rate is temporally invariant. Ref.^37^ correct for this by assuming that the dominant fluctuations in Eq. 16 are caused by variation in the reporting rate *α*_*t*_ rather than in external noise (*u*_*t*_). Eq. 16 can then be expressed as

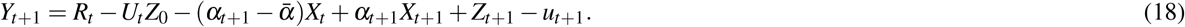

Local linear regression techniques can then be applied to estimate both the reporting rate and the susceptible class. This method is sensitive to the bandwidth parameter, and must be tuned beforehand to minimize large-scale fluctuations from the global mean. We use this ‘locally linear regression method’^37^ for all of the results reported in our paper. The susceptible reconstruction time series from the incidence data in Figure 1 appears in Supplementary Figure 40, and the resulting transmission rate reconstruction time series also appears in Supplementary Figure 41.

### Incidence to Prevalence Conversion

The *prevalence* of the infection is defined by the number (or proportion) of infectious individuals at any given time. Compartmental epidemic models usually predict infection prevalence. However, data are usually in the form of newly occurring cases, referred to as *incidence*. Hence both the proportion of susceptible individuals and the prevalence of infection must be recovered from the infection incidence data before the SINDy algorithm can be applied. Given temporal case incidence data *C*_*t*_, suppose that the duration of infection (*D*_*i*_), the mean individual lifespan (*L*), and the proportion of people that will contract the infection in their lifetime (*p*) are known and constant. The average proportion of the population that is infected at any given time is then given by

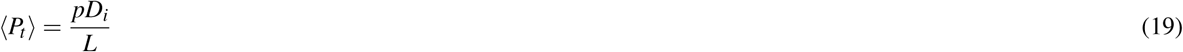

From the relation

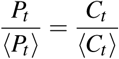

we then obtain

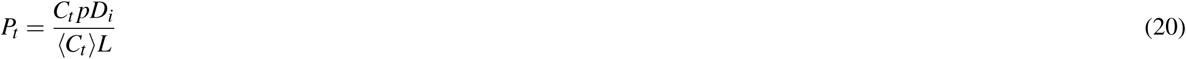

which is used to construct the prevalence (infectious) class given incidence data. We assumed *D*_*i*_ = 2 weeks for all *i*^24,26,76^, *L* = 65 years^77^ and *p* = 0.95^76^.

### Weighted Thresholding

SINDy is based on sparse regression, a statistical learning technique that performs feature selection while fitting the active terms to the data. The realization of this technique used in Ref.^10^ is the iterated thresholding method. The key parameter in this algorithm is *λ*, a chosen threshold below which coefficients (and their corresponding functions) are eliminated on any given iteration. In Ref.^10^ and subsequent papers this parameter is taken as constant, though Ref.^11^ analyses the effects of fitting *λ* using cross-validation. However, epidemiological data present an additional challenge, as the state variables are often orders of magnitude apart (the proportion susceptible, for instance, is *O*(0.1) while the proportion infected is *O*(0.0001). When evaluating a higher order function library using data on contrasting scales, high order functions of small state variables (such as *I*^3^) have a much smaller column norm than larger state variables or functions with a smaller polynomial order. As a result, the iterated sparse regression algorithm can assign them large coefficients to account for this, which are much less likely to be eliminated by a fixed thresholding value.

To account for this, we introduce a threshold for each function in the library that is scaled according to the norm of the corresponding column (see also Ref.^10^). For each column *k* in the function library Θ(*X*), we construct the threshold

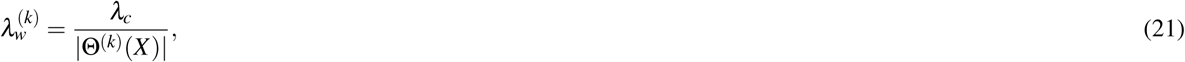

where Θ^(*k*)^(*X*) is the *k*th column in the function library, | · | is the *l*_2_ − norm, and *λ*_*c*_ is a constant threshold value. The algorithm in Section is then performed in the same way, using this function-dependent sparsity knob instead. This is the technique utilized throughout this paper, and any reference to a constant *λ* value is the *λ*_*c*_ parameter in Eq. 21.

### Choice of Functional Basis

Determining the correct basis of elementary functions is a key step when generating a model using SINDy, and the lack of a rigorous method to identify such a basis is one of its notable downfalls^10^. Nevertheless, most compartmental models in epidemiology have been constructed using a simple basis of polynomial and trigonometric functions, which is what we use in this analysis. Many compartmental epidemic models only use polynomial functions on the second degree or lower, so we commonly limit our function library to second or third order polynomials.

Depending on the nature of the system and the assumptions made, it becomes necessary to add several features to the function library. The dynamics of the prevalence of both measles and chickenpox are strongly dictated the seasonal forcing function^24^,^26^. Hence, a new parameter *β* is constructed such that

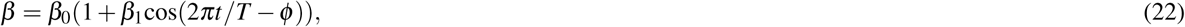

where *T* is the period of the seasonal oscillations (usually 1*yr*^−1^) and *ϕ* is the phase shift. This parameter is then multiplied by each of the *p* columns in Θ to create *p* new features in the function library.

Model dynamics are sensitive to *ϕ*. Hence, we included *ϕ* in a three-dimensional parameter sweep involving *ϕ, S*_0_, and *λ* as follows. For each point of the *S*_0_ − *λ* parameter grid (*λ* ∈ [0.0001, 0.1], *S*_0_ ∈ [0.05, 0.13], see Supplementary Figures 5-12), we also conducted a parameter sweep for *ϕ* ranging from 0 to 52 weeks in increments of Δ*ϕ* = 0.5wk. The SINDy model was generated for each value of *ϕ* in this parameter range, and the SINDy model with lowest AIC values was selected to represent that point in the *S*_0_ − *λ* parameter grid. (We opted for this approach to facilitate visualization of the three-parameter sweep in the two-dimensional *S*_0_ − *λ* plane and to focus on the *S*_0_ − *λ* relationship. Altering the values of *ϕ* away from the optimal values usually worsens the model fit in predictable ways. For instance, a value of *ϕ* that causes transmission to peak in the summer months also causes an epidemic peak in the summer, which is rarely observed in the empirical datasets.)

Given that the susceptible population is influenced heavily by the birth rate, the addition of a birth parameter is also beneficial. A functional form of the birth rate can be assumed and added to the library, but given that in the place and time period of this study the birth rate does not behave in a way that can be described by a linear or exponential function we choose to represent the birth rate in the function library by simply including a column of the empirical data *B*(*t*) that gives the total number of births in week *t*. This data is already required to scale the state variables and the source for each location used in this paper is given in Section.

### Power Spectral Density

Estimates of the power spectral density of the prevalence time series can be useful for model selection when qualitative features such as attractor class are seen as more important than goodness-of-fit. However, a model selection method that values sparsity is still desirable. This leads to a model selection process of computing the AIC score of the spectral densities of the model and the data, which will promote a parsimonious model that attempts to match the qualitative features present in the data. We followed standard procedures for computing power spectra of time series^26^,^78,79^. First, the infection time series data from both the discovered SINDy model and the relevant data were smoothed using a moving average window with a span of 23 timesteps. Second, the smoothed data were linearly trend-corrected. Third, 20 % of the time series was tapered using a split cosine bell. The periodogram of the resulting time series was then computed. We used the Matlab functions smooth, detrend, and periodogram^80^. The AIC score was then computed using the residual sum of squares between the empirical and SINDy model power spectral density estimates, taking the number of free parameters from the discovered model. The methodology for computed the power spectral density described in the out-of-sample prediction subsection was the same.

### SIR Model Fitting

This can be done by simulating the model across a wide range of linearly-spaced parameter values and selecting the model which minimizes the sum of squares error between the simulated model and the observed data. Baseline values for the parameters were taken from Refs.^24^,^76^. For simplicity a closed system was assumed, implying that the birth and death rates were equal (*ν* = *µ*). Also, recall that given basic reproductive ratio *ℛ*_0_ and recovery rate *γ*, the mean transmission rate is completely determined by *β*_0_ = *ℛ*_0_ *γ*. The parameters that were varied, with corresponding ranges and step sizes, were *R*_0_ ∈ (6, 16), *γ ∈* (0.55, 1.25), *β*_1_ *∈* (0.05, 0.35), *µ ∈* (3 × 10^−4^, 6×10^−4^) and (0, 51.5). Simulations of the discrete SIR model were run at each point in the parameter plane, beginning 150 years prior to the temporal range of the data to eliminate the effects of transients and the impact of the initial conditions, which were fixed at (*S*_0_, *I*_0_) = (0.1, 5 ×10^−5^). The resulting models with the minimal sum of squares error when compared with the data were selected and plotted in Supplementary Figures 42-44.

## Data Availability

The code used to generate the results is publicly available at Ref.^62^. The infectious disease data for measles, rubella and chickenpox can be obtained from the International Infectious Disease Data Archive (http://iidda.mcmaster.ca/). Demographic data are available from published sources^68^,70–73.

## Data Availability

The code used to generate the results is publicly available on Github. The infectious disease data for measles, rubella and chickenpox can be obtained from the International Infectious Disease Data Archive.

https://github.com/jonathanhorrocks/SINDy-data

http://iidda.mcmaster.ca/

## Acknowledgments

The authors are grateful to David J.D. Earn for providing data for this project from the International Infectious Disease Data Archive (IIDDA), and to Sri Namachivaya, Giang Tran, and two anonymous reviewers for helpful comments.

## Author contributions statement

CTB conceived the study. CTB and JH developed the methodology. JH conducted the analysis and drafted the manuscript. CTB wrote the final version of the manuscript.

## Additional information

The authors declare no competing interests.

